# Covid-19 serology in nephrology health care workers

**DOI:** 10.1101/2020.07.21.20136218

**Authors:** Thomas Reiter, Sahra Pajenda, Ludwig Wagner, Martina Gaggl, Johanna Atamaniuk, Barbara Holzer, Irene Zimpernik, Daniela Gerges, Katharina Mayer, Christof Aigner, Robert Straßl, Sonja Jansen-Skoupy, Manuela Födinger, Gere Sunder-Plassmann, Alice Schmidt

## Abstract

**Background:** Chronic kidney disease patients show a high mortality in case of a SARS-CoV-2 infection. Thus, to be informed on Nephrology personnel’s sero-status might be crucial for patient protection. However, limited information exists about the presence of SARS-CoV-2 antibodies in asymptomatic individuals.

**Methods:** We examined the seroprevalence of SARS-CoV-2 IgG and IgM antibodies among health care workers of a tertiary care kidney center during the peak phase of the Covid-19 crisis in Austria using an orthogonal test strategy and a total of 12 commercial nucleocapsid protein or spike glycoprotein based assays as well as Western blotting and a neutralization assay.

**Results:** At baseline 60 of 235 study participants (25.5%, 95% CI: 20.4-31.5) were judged to be borderline positive or positive for IgM or IgG using a high sensitivity/low specificity threshold in one test system. Follow-up analysis after about two weeks revealed IgG positivity in 12 (5.1%, 95% CI: 2.9-8.8) and IgM positivity in six (2.6%, 95% CI: 1.1-5.6) in at least one assay. 2.1% (95% CI: 0.8-5.0) of health care workers showed IgG nucleocapsid antibodies in at least two assays. By contrast, positive controls with proven Covid-19 showed antibody positivity among almost all test systems. Moreover, serum samples obtained from health care workers did not show SARS-CoV-2 neutralizing capacity, in contrast to positive controls.

**Conclusions:** Using a broad spectrum of antibody tests the present study revealed inconsistent results for SARS-CoV-2 seroprevalence among asymptomatic individuals, while this was not the case among Covid-19 patients.

**Trial registration number:** CONEC, ClinicalTrials.gov number NCT04347694

## Introduction

Health care workers are at increased risk for severe acute respiratory syndrome-coronavirus-2 (SARS-CoV-2) infection resulting in severe coronavirus disease 2019 (Covid-19).^1-3^ Properly used protection equipment can reduce transmission risk, but direct patient contact, endotracheal intubation, or contact with contagious body fluids are associated with an increased infection risk.^4^ In turn, infected health care workers pose a significant threat to patients they care for.^5^ People with a compromised immune system or on treatment with immunosuppressive drugs, such as patients with chronic kidney disease (CKD) including those on dialysis treatment or with a kidney transplant, are among the most vulnerable with respect to life threatening infectious diseases.^6-8^ Regardless of the “stay at home-stay safe” practice during the Covid-19 pandemic, they are in need of non-deferrable admissions to kidney centers.

Reports on SARS-CoV-2 infection among patients with CKD showed a mortality of up to 28% in kidney transplant recipients or solid organ transplants.^9-11^ Early studies from China revealed a surprisingly low mortality in dialysis patients, which contrasts with reports from the Austrian Dialysis and Transplant registry.^12,13^ On May 8^th^ 2020 Covid-19 specific mortality was 27% (12/44), which is comparable to the reported rate of 31% (18/59) in a recent report from the Columbia University Irving Medical Center, NY.^14,15^ It is well established that containment strategies in Austria were successful in preventing a collapse of the acute care facilities. More importantly, Covid-19-specific mortality was low as compared to other European countries.^16,17^

While reverse transcriptase-polymerase chain reaction (RT-PCR) amplification of SARS-CoV-2 RNA has its utility to identify acutely infected patients, serology testing is important to identify patients that have been infected in the past. Thus, detection of SARS-CoV-2 specific antibodies is a prevalence marker in a population and can be used to measure herd immunity.^18-20^ In patients suffering from Covid-19 with RT-PCR proven SARS-CoV-2 infection up to 100 % tested positive for antiviral immunoglobulin-G (IgG) within three weeks after symptom onset. Seroconversion for IgG and immunoglobulin-M (IgM) occurred simultaneously or sequentially.^21-25^ However, the antibody response to SARS-CoV-2 and seroprevalence among asymptomatic health care workers is far from clear. In this regard, the SARS-CoV-2 immune status of personnel of kidney centers is of eminent importance due to the susceptibility of renal patients to Covid-19.

We aimed to examine the prevalence of SARS-CoV-2 antibodies among nephrology healthcare workers in a tertiary care, university-based hospital in Austria. We adhered to an orthogonal test strategy and used a comprehensive set of commercial laboratory tests and Western blotting, including Covid-19 controls and analysis of neutralizing antibodies.^26^

## Methods

### Study design and participants

The “COvid-19 Serology in NEphrology Health Care Workers” (CONEC, ClinicalTrials.gov no. NCT04347694) study is a longitudinal study examining the antibody response to SARS-CoV-2 among staff members of the Division of Nephrology and Dialysis, Department of Medicine III, at the Medical University of Vienna, Austria. We enrolled nurses, doctors, researchers administrators, cleaners, and other staff. The study protocol includes sample collection at baseline and every three months thereafter for a minimum of one year. Serum samples were stored at -80°C before testing. At each study visit, participants filled out a questionnaire including demographic data, job title, medical history, medication, travel history since the beginning of the Covid-19 crisis in Austria, Covid-19 specific history including contact to proven Covid-19 cases, Covid-19 specific symptoms, and results of non-study-related SARS-CoV-2 laboratory tests. At baseline, participants were screened for the presence of serum anti-SARS-CoV-2 IgG and IgM antibodies by means of the ImmunoDiagnostics test system (ImmunoDiagnostics, HongKong).^27^ Subjects with a borderline positive or positive initial antibody test were invited to follow-up serum antibody tests, and a SARS-CoV-2 RT-PCR, within two to four weeks. Employing this extended orthogonal test strategy, the ImmunoDiagnostics test was repeated and samples were also tested by a set of other commercial laboratory tests, covering nucleocapsid protein and spike glycoprotein specific tests for anti-SARS-CoV-2-IgG, -IgM, and –IgA, as well as by nucleocapsid protein and spike glycoprotein Western blots.^26^ We used two additional anti-SARS-CoV-2 IgG ELISA tests and a microscopy-based neutralization assay with authentic SARS-CoV-2 to confirm the presence of SARS-CoV-2 specific neutralizing or non-neutralizing antibodies in all follow-up samples that previously tested positive for any anti-SARS-CoV-2 IgG antibodies. Serum samples of five Covid-19 patients served as positive controls for all follow-up laboratory tests. We report here the baseline data of the CONEC study, which was approved by the institutional review board (IRB) at the Medical University of Vienna (unique IRB identifier: 1357/2020). All participants provided written informed consent.

### Laboratory analysis

We used a set of ten commercial serologic tests for detection of anti SARS-CoV-2 IgG (two for antigenic target nucleocapsid protein, two for antigenic target spike glycoprotein), of anti-SARS-CoV-2 IgM (two antigenic target nucleocapsid protein, one antigenic target spike glycoprotein), of anti-SARS-CoV-2 IgA (one antigenic target spike glycoprotein), and of anti-SARS-CoV-2 total antibody (one antigenic target nucleocapsid protein, one antigenic target spike glycoprotein) including enzyme-linked immunosorbent assays (ELISA), chemiluminescence immunoassay (CLIA), and electrochemiluminescence immunoassay (ECLIA), according to the instructions of the manufacturers, for all follow-up analyses **(Supplementary appendix)**. Technical details of these tests are indicated in **Supplementary Table S1**. In subjects with IgG antibodies at follow-up and for Covid-19 control samples we also used two additional IgG ELISAs. Western blotting, analysis of neutralizing antibodies, and SARS-CoV-2 RT-PCR are described in the **Supplementary appendix**.

### Statistical analysis

Demographic information at baseline and follow-up are given in means (standard deviation) and count (percent), respectively. For the main outcome (occult immunization yes/no), results are tabulated for each time point. All prevalence estimates are presented with 95% confidence intervals approximated by the Wald method. Formal statistical testing for categorical data is done by the Fisher Exact test. Continuous data is analyzed by unpaired t-tests or by non-parametric tests in cases of non-normally distributed data. Data management and statistical analysis has been performed by means of MS Excel (Microsoft, Redmond, WA) and R (R Core Team (2016), R Foundation for Statistical Computing, Vienna, Austria).

## Results

### Participants

Beginning four weeks after the first documented Covid-19 cases in the Austrian states of Tyrol and Vienna in late February 2020, and two weeks after the shutdown in our country, we approached 288 health care workers at the Division of Nephrology and Dialysis at the Medical University of Vienna for participation in this study. The different sections of care of our Department as well as the number and occupations of study participants are indicated in **Supplementary Figure S1** and **S2**. A total of 235 staff members (82%) agreed to enter the study and had their first study visit during a period of about four weeks. The rate of participation was 94.4% among physicians, 74.4% among nurses, 90.9% among researchers, 84.6% among administrative staff and 89.2% among other staff.

Demographic and general clinical data of all participants and Covid-19 related history of these individuals is given in **Table 1**. Eleven (4.7%) participants reported contact with proven Covid-19 cases, and 36 (15.3%) with category 1 or 2 persons. A travel history outside or inside Austria was evident in 93 (39.6%) cases. Two (0.9%) were in quarantine and none reported a history of Covid-19 or home isolation at baseline. However, 62 (26.4%) persons reported symptoms possibly related to undetected mild Covid-19 (**Table 1**). Nineteen (8.1%) of these subjects reported a previous SARS-CoV-2 RT-PCR at baseline, which was negative in all cases. During the study period, a total of 313 nasopharyngeal swabs were negative in 179 (76.2%) participants (56 participants had no SARS-CoV-2 RT-PCR).

**Table 1.** Demographic characteristics and Covid-19 related history.

| Characteristic | N=235 |
| --- | --- |
| Age, years (mean ±SD) | 44.2±11.4 |
| Sex, female, no. (%) | 165 (70.2%) |
| BMI, kg/m<sup>2</sup> (mean ±SD) | 25.4±4.7 |
| Profession, no. (%) |  |
| - Physician | 51 (21.7%) |
| - Nurse | 119 (50.6%) |
| - Researcher | 10 (4.2%) |
| - Administrative staff | 22 (9.4%) |
| - Other staff | 33 (14.0%) |
| Comorbidities, no. (%) |  |
| - Hypertension | 36 (15.3) |
| - Diabetes | 6 (2.6) |
| - Coronary artery disease | 4 (1.7) |
| - Chronic kidney disease | 2 (0.85) |
| - Lung disease | 16 (6.8) |
| - Autoimmune disease | 18 (7.7) |
| - Cancer | 5 (2.1) |
| Clinical symptoms, no. (%) |  |
| - Fever | 10 (4.2) |
| - Cough | 34 (14.5) |
| - Dyspnea | 4 (1.7) |
| - Gastrointestinal complaints | 18 (7.7) |
| - Loss of smell and taste | 7 (3.0) |
| - Other complaints | 27 (11.5) |
| Smoking history, no. (%) |  |
| - Never smoked | 128 (54.5) |
| - Former smoker | 53 (22.6) |
| - Current smoker | 53 (22.6) |
| ACEI or ARB use, no. (%) | 21 (8.9) |
| History of influenza vaccination, no. (%) | 106 (45.1) |
| Travel to other countries since February 2020, no. (%) | 43 (18.3) |
| Travel within Austria since February 2020, no. (%) | 50 (21.3) |
| SARS-CoV-2 RT-PCR before study entry, no. (%) | 19 (8.1) |
| - Positive | 0 |
| Subjects in same household, median (IQR) | 1 (1-2) |
| - SARS-CoV-2 RT-PCR positive in same household, no. (%) | 0 |
| Contact to Covid-19 patients before study entry, no. (%) | 11 (4.7) |
| Contact to category 1 or 2 individuals before study entry, no. (%) | 36 (15.3) |

Covid-19, coronavirus disease 2019; BMI, body mass index; SD, standard deviation; ACEI, angiotensin converting enzyme inhibitor; ARB, angiotensin receptor blocker; SARS-CoV-2, severe acute respiratory syndrome-coronavirus-2; RT-PCR, reverse transcriptase-polymerase chain reaction; IQR, inter quartile range.

### SARS-CoV-2 antibodies - *baseline*

Among 235 participants, we judged 60 (25.5%, 95% CI: 20.4-31.5) at baseline to be either borderline positive or positive for anti-SARS-CoV-2 IgG and/or IgM by the ImmunoDiagnostics ELISAs using a conservative threshold of an OD of 0.200 and 0.300 in at least one of two duplicates, respectively (Figure 1). Thus, 18 (7.7%, 95% CI: 4.8-11.9) individuals were assumed to be IgM positive, 3 others (1.3%, 95% CI: 0.03-3.9) IgG positive, and the remaining 39 (16.6%, 95% CI: 12.4-21.9) borderline positive for IgM and/or IgG.

**Figure 1.**
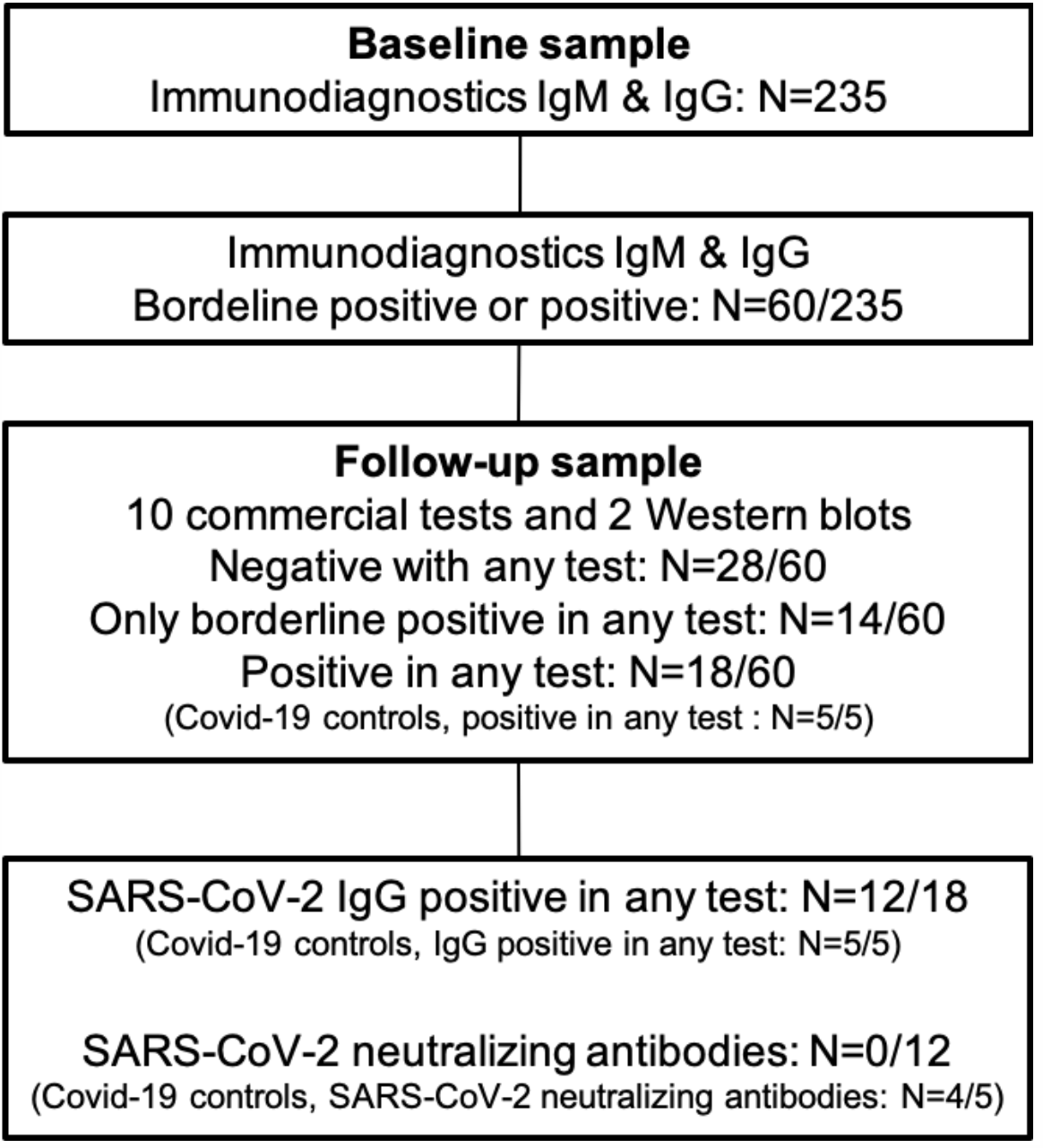
Overview of main results.

### SARS-CoV-2 antibodies - *follow-up*

All 60 follow-up participants had a serum test and the majority a SARS-CoV-2 RT-PCR within 18.5±7.0 days after baseline. Their clinical characteristics are indicated in **Table 2**. All but five, who had no RT-PCR, tested negative for SARS-CoV-2 RNA in nasopharyngeal swabs at this point in time. Details of test results of 28 (46.7%, 95% CI: 34.6-59.1) follow-up negative and 14 (23.3%, 95% CI: 14.3-35.6) borderline positive participants (**Figure 1**) are given in the **Supplementary appendix**.

**Table 2.** Baseline and follow-up clinical characteristics and Covid-19 related history of 60 individuals with a borderline positive or positive SARS-CoV-2-IgG and/or -IgM ImmunoDiagnostics test at study entry, and of subgroups comprising 42 who tested negative or borderline positive at follow-up in any test system (subgroup 1), and of 18 with a positive test result in any test system at follow-up (subgroup 2).

| Characteristic | All follow-up<br>N=60<sup>a</sup> |  | Subgroup 1<br>N=42<sup>b</sup> |  | Subgroup 2<br>N=18<sup>c</sup> |  | P | P |
| --- | --- | --- | --- | --- | --- | --- | --- | --- |
|  | BL | FU<sup>d</sup> | BL | FU<sup>d</sup> | BL | FU<sup>d</sup> | Between BL | Between FU |
| Age, years (±SD) | 41.7 (12.6) | - | 40.6 (12.9) | - | 44.4 (11.7) | - | 0.2936 | - |
| Female sex, no. (%) | 49 (81.7) | - | 37 (88.1) | - | 12 (66.7) | - | 0.07 | - |
| History of influenza vaccination, no. (%) | 23 (38.3) | - | 17 (40.5) | - | 6 (33.3) | - | 0.7734 | - |
| History of autoimmune disease, no. (%) | 6 (10.0) | - | 4 (9.5) | - | 2 (11.1) | - | 1 | - |
| Smoker, no (%) | 13 (21.7) | - | 6 (14.3) | - | 7 (38.9) | - | 0.04561 | - |
| Days between BL and FU (±SD) | - | 18.5 (7.1) | - | 18.8 (7.5) | - | 17.6 (5.7) | - | 0.4771 |
| Clinical symptoms, no. (%) |  |  |  |  |  |  |  |  |
| - Fever | 1 (1.7) | 0 | 0 | 0 | 1 (5.6) | 0 | 1 | 1 |
| - Cough | 12 (20.0) | 5 (8.3) | 7 (16.7) | 3 (7.1) | 5 (27.8) | 2 (11.1) | 0.4819 | 0.6311 |
| - Dyspnea | 2 (3.3) | 1 (1.7) | 2 (4.8) | 1 (2.4) | 0 | 0 | 1 | 1 |
| - Gastrointestinal complaints | 4 (6.7) | 2 (3.3) | 2 (4.8) | 2 (4.8) | 2 (11.1) | 0 | 0.5762 | 1 |
| - Loss of smell and taste | 4 (6.7) | 1 (1.7) | 3 (7.1) | 1 (2.4) | 1 (5.6) | 0 | 1 | 1 |
| Travel to other countries, no. (%) | 10 (16.7) | 0 | 8 (19.0) | 0 | 2 (11.1) | 0 | 0.708 | - |
| Travel within Austria, no. (%) | 14 (23.3) | 8 (13.3) | 8 (19.0) | 4 (9.5) | 6 (33.3) | 4 (22.2) | 0.3188 | 0.2252 |
| Subjects in same household, median (IQR) | 1 (1-2) | 1 (1-2) | 2 (1-2) | 1.5 (1-2) | 1 (1-2) | 1 (1-2) | 0.2943 | 0.4031 |
| - SARS-CoV-2 RT-PCR positive, no. (%) | 0 | 0 | 0 | 0 | 0 | 0 | - | - |
| Contact to Covid-19 patients, no. (%) | 4 (6.7) | 0 | 2 (4.8) | 0 | 2 (11.1) | 0 | 0.5762 | - |
| Contact to category 1 or 2 individuals, no. (%) | 7 (11.7) | 0 | 5 (11.9) | 0 | 2 (11.1) | 0 | 1 | - |

Covid-19, coronavirus disease 2019; SARS-CoV-2, severe acute respiratory syndrome-coronavirus-2; BL, baseline; FU, follow-up; SD, standard deviation; RT-PCR, reverse transcriptase-polymerase chain reaction; IQR, inter quartile range.
<sup>a</sup>, physicians , 12 (20%), nurses, 28 (46.7%), research staff, 2 (3.3%), administration, 5 (8.3%), other staff, 13 (21.7%).
<sup>b</sup>, physicians , 8 (19.0%), nurses, 19 (45.2 %), research staff, 2 (4.8 %), administration, 3 (7.1 %), other staff, 10 (23.8 %).
<sup>c</sup>, physicians, 4 (22.2%), nurses, 9 (50%), research staff, none, administration, 2 (11.1%), other staff, 3 (16.7%).
<sup>d</sup>, numbers refer to time period between baseline and follow-up.

Eighteen follow-up participants - 30.0% (95% CI: 19.8-42.6) or 7.7% (95% CI: 4.8-11.9) of the total cohort - showed a positive SARS-CoV-2 antibody result in one or more laboratory tests at follow-up (**Figure 1** and **Figure 2**). Details of these 18 participants are indicated in **Table 2**. All clinical characteristics but smoking (OR: 3.82, 95% CI: 1.06-13.77, p<0.05, for presence of antibodies for smokers) did not differ between follow-up positive as compared to follow-up negative or borderline positive participants at baseline or follow-up. Individual test results of these 18 subjects are shown in **Supplementary Table S2 and S3** and explained in the **Supplementary appendix**.

**Figure 2.**
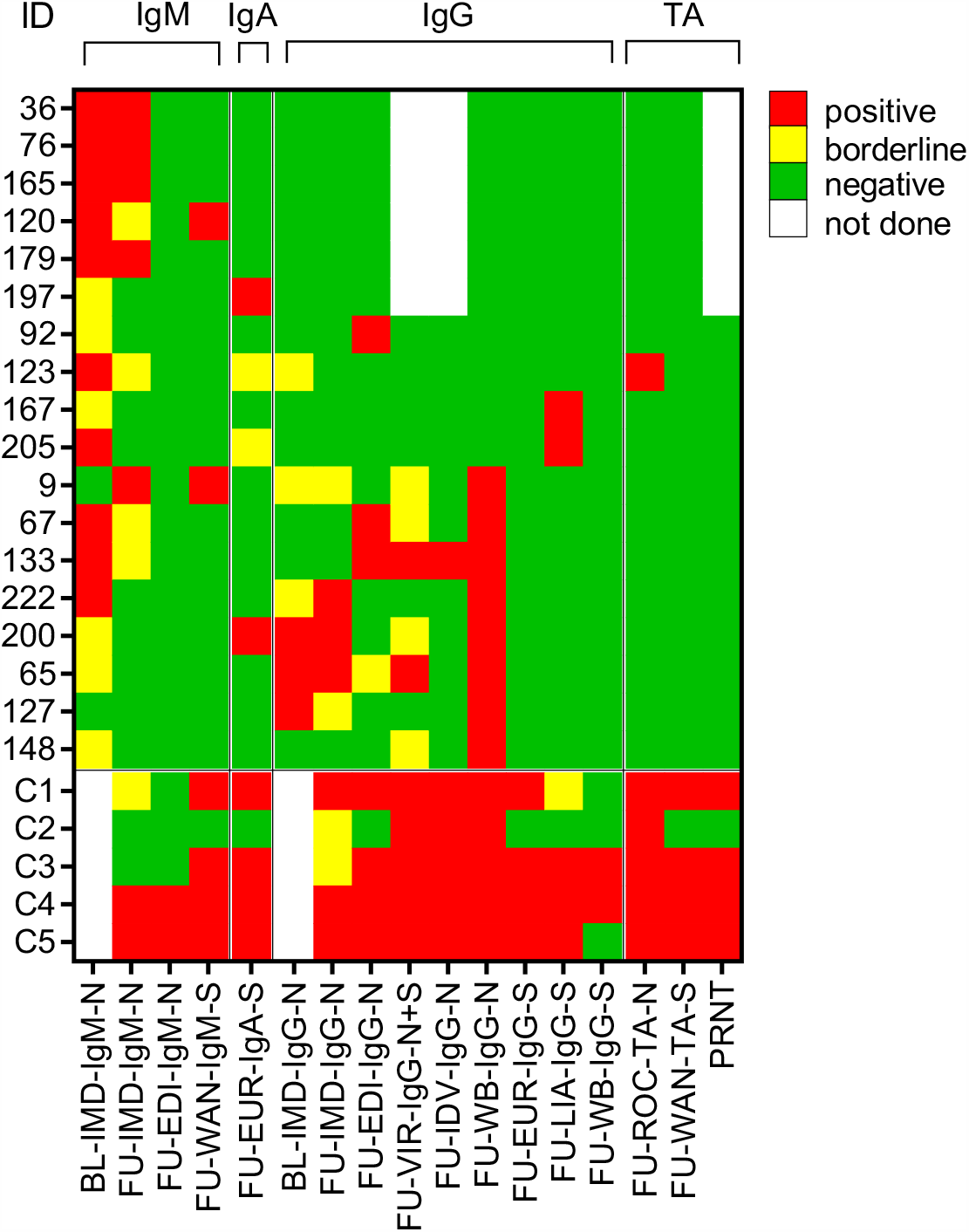
Baseline and follow-up SARS-CoV-2 antibody test results of 18 study participants positive in at least one test system at follow-up and of five Covid-19 patients. The laboratory results of five Covid-19 patients, indicated by C1 to C5, are shown in the bottom lines corresponding to the follow-up test results of study participants. TA, total antibody; BL, baseline; IMD ImmunoDiagnostics, HongKong; N, SARS-CoV-2 nucleocapsid protein; FU, follow-up; EDI, Epitope Diagnostics Inc., San Diego, CA; WAN, Beijing Wantai Biological Pharmacy Enterprise Co., Ltd., Bejing, China; S, SARS-CoV-2 spike glycoprotein; EUR, Euroimmun Medizinische Labordiagnostika AG, Lübeck, Germany; VIR, Vircell, Granada, Spain; IDV, IDvet, Grabels, France; WB, Western Blot; LIA, Liaison, DiaSorin S.p.A, Saluggia, Italy; ROC, Roche Diagnostics Deutschland G.m.b.H, Mannheim, Germany; PRNT, plaque reduction neutralization test.

Six participants (2.6%, 95% CI: 1.1-5.6) showed IgM and two (0.9%, 95% CI: 0.03-3.3) showed IgA antibodies at follow-up. With regard to anti-SARS-CoV-2 IgG, 10 of 18 individuals had anti-nucleocapsid IgG and two had anti-spike glycoprotein IgG in at least one test system, resulting in an overall prevalence of anti-SARS-CoV-2 IgG of 5.1% (12/235; 95% CI: 2.9-8.8%.) None of these 12 study participants had neutralizing SARS-CoV-2 antibodies, but two of them also tested positive for IgG by the Vircell and the IDvet ELISAs (**Figure 2**). At least two positive IgG antibody results were seen in five of 18 subjects (2.1% of the entire study cohort, 95% CI: 0.7-5.0; all anti-nucleocapsid protein IgG; ID 65, 67, 133, 200, and 222 in **Figure 2**, and **Supplementary Table S2** and **S3**). Only one of these individuals reported symptoms possibly related to mild Covid-19 (ID 133). The proportion of SARS-CoV-2 antibody positive subjects among the different professions is shown in **Supplementary Figure S5**.

All five Covid-19 serum samples (case vignettes can be found in the **Supplementary appendix**) tested positive for SARS-CoV-2 antibodies in the majority of tests (mean time between first positive SARS-CoV-2 RT-PCR and serum sampling: 31.8±12.2 days). Four out of five patients had neutralizing SARS-CoV-2 antibodies. The other one had the mildest form of Covid-19 among positive controls presenting with joint pain (**Figure 2, Supplementary appendix, Supplementary Table S2** and **S3**).

## Discussion

Our study addressess several important issues related to Covid-19: First, the seroprevalence of anti-SARS-CoV-2 IgG antibodies among nephrology healthcare workers at the Medical University of Vienna during the infection peak in Austria was, at its best, 2.1% (95% CI: 0.7-5.0). Second, it is valid to assume that the successful containment measures taken in Austria and especially in Vienna have minimized exposure of health care workers at our institution to Covid-19 in the community and at the point of care as compared to other countries. Third, commercially available laboratory tests, including ELISA, CLIA, and ECLIA, may fail to uniformly detect potential low-level immune response to SARS-CoV-2 in asymptomatic subjects or mild disease, or differentially cross-react as false-positives.

The analytical specificity of a laboratory test is reflected by the positive predictive value (PPV), which depends not only on the sensitivity and specificity of the test but also on the disease prevalence. Requiring a PPV of at least 90%, the analytical specificity of a test should ideally exceed 99.9%, which is influenced by the presence of autoimmune diseases, heterophilic antibodies, or antibodies to other coronaviruses.^28,29^ At follow-up, we found no effect of influenza vaccination or history of autoimmune disease on Covid-19 serostatus. However, there were more smokers among antibody positives. This finding may be related to the preference of nicotine for the ACE2-SARS-CoV-2 complex that reduces SARS-CoV-2 virulence by interference with the spike protein.^30^

Overall, serologic tests based on spike glycoprotein appear to distinguish between emerging and endemic coronaviruses, whereas assays based on the nucleocapsid protein can serve as a marker of recent infection but might be expected to cross-react more with endemic coronaviruses.^31^ Test reactivity thresholds used to define a positive result can be adjusted to optimize the tradeoff between sensitivity and specificity. With higher thresholds, sensitivity decreases as cases with low serum antibody levels are categorized as negative, but specificity improves, as low amounts of nonspecific antibody are no longer considered positive.^31^ In our study, we used a low threshold for IgM and IgG ELISAs at baseline to account for higher sensitivity, accepting low specificity as demonstrated by follow-up examinations.

A meta-analysis of 38 studies covering 7,848 individuals confirmed that tests using the spike glycoprotein are more sensitive than nucleocapsid-based tests. IgG tests performed better compared to IgM tests with higher sensitivity at later time points after the onset of symptoms. Combined IgG and IgM tests performed better in terms of sensitivity than measuring either antibody alone. All methods yielded high specificity with some tests reaching levels around 99%.^32^ However, statistically the PPV varies widely and can be as low as 30% to 50% in low prevalence settings.^33^

Rigorous containment strategies may also reduce the prevalence of anti-SARS-CoV-2 in different populations at the cost of an early development of herd immunity. Travel restrictions and other control measures reduced Covid-19 transmission early this year in China.^34^ Later, it was estimated that, among 11 European countries, the national lockdown had the greatest effect on the reproduction number R_t_ among non-pharmacologic interventions including school closure, avoidance of public events, social distancing, and self-isolation. As such, Austria (**Supplementary Figure S6**) and Norway had the lowest infection rate in this analysis.^17^ At the beginning of this study the R_t_ in Vienna was 2.0 (95% CI: 1.87-2.14) and further decreased thereafter. The implementation of public interventions affects the case number after about two weeks, and employment of econometric techniques showed that policy changes in six countries across the globe averted 530 million infections.^35,36^

The low seroprevalence of Covid-19 antibodies in nephrology health care workers in our institution reflects successful measures taken to prevent transmission/infection by the city of Vienna and the Medical University of Vienna. Taken together, this minimized exposure risk to Covid-19 for our staff at work (**Supplementary Figure S7** and **S8**).

The prevalence of asymptomatic cases among SARS-Cov2 infected patients is supposed to be 40-45%.^37^ In our cohort one of five participants considered to be IgG positive showed symptoms potentially related to SARS-CoV-2 exposure. In contrast, significant exposure to Covid-19 cases resulted in a seroprevalence of 17.4% and 44% among health care workers in the US and in China, respectively.^38,39^ Other studies in high-risk settings, however, showed a low seroprevalence among hospital staff.^40-44^ These surveys utilized only one test system and none of these studies employed an orthogonal strategy, confirming borderline positive or positive samples in an independent follow-up serum sample using other laboratory tests.

The extended orthogonal test strategy of the present study, including 12 different commercial tests, Western blots, and a neutralization test, has potentially allowed for an increase in sensitivity/specificity for confirmation of seropositivity among some individuals. Assuming true seropositivity in five of 18 health care workers, with positive IgG titers in at least two of the commercial or in-house test systems at follow-up (uniform nucleocapsid protein IgG in all five cases, also pointing to potential cross-reaction), suggests that single antibody tests do not allow for correct detection of true seroconversion and have no acceptable sensitivity and/or specificity in largely asymptomatic and SARS-CoV-2 RT-PCR negative individuals. This is nicely shown by the rag-rug pattern of seroconversion in **Figure 2**. Of note, non-exposed healthy subjects harbor pre-existing SARS-CoV-2 cross-reactive T cells, specific for a huge array of SARS-CoV-2 antigens, suggesting some potential for pre-existing immunity in the population.^45^ This finding matches, at least in part, with the presence of SARS-CoV-2 antibodies in non-exposed individuals.

In contrast, all five Covid-19 serum samples in our study showed SARS-CoV-2 IgM and IgG antibodies in more than two test systems (**Figure 2**). Four had neutralizing antibodies, whereas none of the serum samples of the IgG positive study participants showed SARS-CoV-2 neutralizing capacity (**Figure 2, Supplementary Table S3**). The differential development of antibody response to nucleocapsid protein and spike glycoprotein antigens among patients with proven Covid-19 and the study participants is in support of more cross-reaction than anti-SARS-CoV-2 seroconversion in health care workers enrolled in this study. A potential limitation to this study relies in the lack of unambiguous Covid-19 cases among study participants, which was not expected to be the case at the beginning of this study in March 2020. We also did not do a formal laboratory test performance analysis. This is largely counterbalanced by the strength of this study, namely the orthogonal test strategy with confirmation in separate serum samples using a very broad range of SARS-CoV-2 antibody tests.

In summary, our study demonstrates that single antibody tests are not reliable to assess the SARS-CoV-2 immune response in mostly asymptomatic individuals. This finding has important implications for testing cohorts with a low Covid-19 prevalence to determine whether herd immunity has been reached. Containment strategies by the City of Vienna and the Medical University of Vienna proved to be extremely effective given the very low seroprevalence in a cohort of high-risk health care workers during the peak of the pandemic crisis in Austria. However, caution has still to be taken, since health care workers are prone to Covid-19 infections and transmission to patients.

## Data Availability

De-identified data will be available through the corresponding author.

## Acknowledgements

We are indebted to Miriam Kunst, Karin Voith, and Gabriele Holzmann for support in sample collection and data management.

